# Engagement With a Breath-Based Metabolic Device Is Associated with Greater Weight Loss in Self-Reported Real-World GLP-1RA Users

**DOI:** 10.64898/2026.02.22.26346841

**Authors:** Gil Ben David, Ronald Udasin, Dan Golan, Michal Mor, Merav Mor

## Abstract

**Background:** Digital health self-monitoring tools are widely used to support weight management and metabolic health. Higher engagement with these tools is often associated with better clinical outcomes; however, real-world engagement-outcome relationships for consumer metabolic monitoring devices remain incompletely characterized, particularly in heterogeneous user populations.

**Objective:** To evaluate whether engagement with a portable breath-based metabolic device (Lumen; Metaflow Ltd.) is associated with greater weight loss and reduction in body fat among real-world glucagon-like peptide-1 receptor agonist (GLP-1RA) users. The study also explores correlations between engagement and a device-specific measure of metabolic flexibility (FLEX score).

**Methods:** We retrospectively analyzed 2,296 adult Lumen users who self-reported GLP-1RA use over 24 weeks. Engagement was quantified as total engagement days over a 24-week period and ordered engagement consistency groups defined by weekly use frequency thresholds. Weight and body fat percentage data were collected by a combination of connected devices and manual user input in the Lumen smartphone application. Associations with weight loss and reduction in body fat percentage were evaluated using linear regression and ANCOVA adjusted for age, baseline BMI, and sex, with HC3 robust standard errors. Body fat percentage data were available for only 490 of the 2,296 subjects. In addition, similar associations were evaluated for FLEX score. GLP-1RA exposure was self-reported at onboarding and not verified longitudinally.

**Results:** At 24 weeks, low/medium/high engagement users lost 3.2%, 4.6%, and 5.2% of body weight (trend p=2.36×10^−^11). Engagement days were associated with percent weight change (slope −0.0214% per day; *P*(HC3)=7.9×10^− 18^). Engagement days showed modest association with body fat percentage change (n=490; slope −0.0105% per day; *P*(HC3)=.010). The adjusted ANCOVA trend across engagement groups was not significant (*P*=.19). Engagement days and consistency both showed a highly significant trend in increase in FLEX score (slope +0.0185 per day; *P*(HC3)=2.0×10^− 36^).

**Conclusions:** In a real-world digital health dataset, higher engagement with a breath-based metabolic monitoring device and its smartphone application was associated with greater 24-week weight loss after adjustment for age, baseline BMI, and sex. The absolute difference between low and high engagement (2.0% body weight) is modest but clinically meaningful in real-world settings after 24 weeks of tracking. Associations with body fat percentage change were smaller and not consistently significant in adjusted analyses. Associations with metabolic flexibility were highly significant, but it remains unknown whether this parameter is predictive or reflective. Prospective controlled studies are needed to test causality and determine whether device-driven biofeedback and sustained engagement independently improve outcomes because GLP-1RA use was self-reported and unverified, and the present analysis was observational. These findings should be interpreted as engagement–outcome associations and reflect behavioral motivation and adherence rather than evidence of device efficacy.

## Introduction

### Background

Obesity and metabolic syndrome are among the leading preventable causes of death worldwide, leading to physiological conditions such as cardiovascular diseases, type 2 diabetes, various types of cancer, as well as social and psychological complications such as depression. In 2022, obesity affected over a billion people worldwide, with prevalence having more than doubled since 1990 [1,2]. The number of annual deaths attributable to high body mass index (BMI) is approaching 4 million [3].

We hypothesized that real-world combined use of a portable breath-based metabolic device that infers fuel utilization from exhaled carbon dioxide percentage with its smartphone application might be associated with improved clinical outcomes related to weight loss and decrease in body fat percentage. The device, Lumen, and its smartphone application comprise a system where every breath a user takes generates data. Then, the application, which integrates this data with that from additional inputs such as fitness trackers, provides personalized insights such as nutritional composition and lifestyle behavioral recommendations. It has already been validated to detect directional shifts in fuel utilization via orthogonal comparison with gold-standard indirect calorimetry [4] and produces expected results in response to meal composition [5]. This is also reflected in both real-world usage and controlled studies [6–8].

Another parameter output from the device is a measure of metabolic flexibility. Metabolic flexibility, the ability to shift substrate oxidation (e.g. whether the primary energy source is lipids or carbohydrates) appropriately as physiological conditions change, such as fasting state and whether a person is in a state of exercise or rest, is vital for maintaining metabolic health and athletic performance [9,10]. Individuals with high metabolic flexibility effectively manage their energy resources, supporting sustained physical and mental functions. In contrast, metabolic flexibility is impaired in and associated with the pathophysiology of a variety of metabolic disorders, including type 2 diabetes, cardiovascular diseases, obesity, and related metabolic and inflammatory conditions. However, literature suggests that metabolic flexibility is modifiable by weight loss and exercise [11–13].

However, the metabolic cart conventionally required to measure metabolic flexibility is expensive and inaccessible, requiring a sophisticated health care laboratory [14]. Because the Lumen system provides breath-based feedback intended to reflect fuel utilization, we evaluated a device-derived metabolic flexibility metric (FLEX score). Additionally, we tested whether engagement with the Lumen system influences FLEX score. The clinical validity of device-derived metabolic flexibility metrics outside laboratory conditions remains uncertain.

We tested this hypothesis in a self-reported cohort of subjects taking glucagon-like peptide-1 receptor agonists (GLP-1RAs). GLP-1RAs have been established to offer benefits in glycemic control, weight reduction through their ability to stimulate insulin secretion and suppress glucagon release, thereby reducing appetite [15,16]. Across multiple randomized clinical trials, median weight loss after 56-72 weeks ranged from 8% to 21% from the GLP-1RAs liraglutide, semaglutide, and tirzepatide [17–19].

A major limitation of GLP-1RA therapy is discontinuation, with up to 65% of patients stopping therapy within one year of initiation [20]. This is attributed to both high cost, lack of access, and side effects such as vomiting, nausea, and diarrhea, abdominal pain, and depression [20–22]. Although GLP-1RA is designed to continue indefinitely, this is often not the case in practice. Therefore, both adherence to drug treatment, as well as continuation of a healthy lifestyle after stopping treatment, could be improved by pairing GLP-1RA use with various types of smartphone applications and dedicated devices that may aid in weight loss. Examples of types of applications already studied as it relates to engagement and weight loss are coaching apps and fitness trackers. For example, a retrospective study in the UK showed that “engaged” users of an application with diet coaching and weight tracking lost 11.5% of body weight after 5 months versus 8.0% for “non-engaged” users [23]. The same group also showed that these results were sustainable at 11 months, where engaged users lost 21.5%, and non-engaged users lost only 17.0% [24]. A different study in Denmark showed that engagement in a behavioral program improved weight loss at 64 weeks from 13.7% to 17.0% [25].

Although GLP-1RA therapy results in weight loss, lifestyle interventions, such as intense behavioral therapy (IBT), are almost always paired with their use to optimize and sustain outcomes. Lifestyle interventions, such as diet and exercise, remain the foundation for obesity treatment, and pharmacotherapy is an important tool to augment their efficacy and durability. A single-site, open-label IBT pilot trial showed 6.1% weight loss after one year with IBT alone versus 11.5% weight loss at one year for IBT + 3 mg liraglutide [26]. A multisite, randomized, double-blind, placebo-controlled study confirmed these findings, showing IBT combined with liraglutide 3.0 mg achieved 7.5% 56-week weight loss compared with 4.0% weight reduction following IBT with placebo [27].

Another area of concern for patients treated with GLP-1RA therapy is that the composition of weight loss can include a meaningful reduction in lean mass (skeletal muscle, body water, and organ mass) in addition to fat loss. Studies have shown that while the majority of the weight loss is indeed from fat, approximately 25% of weight loss is from lean mass [28,29]. Current mitigation strategies are focused on nutritional guidance emphasizing adequate protein intake and resistance training in order to maintain and build muscle mass, especially in older individuals who are at greater risk for sarcopenia [30–32]. As we do not have a direct measure of lean mass, we use estimated body fat percentage to represent this parameter in the current study.

### Objective

In the present study, we investigated whether level of engagement with the Lumen system (device and smartphone application) is positively correlated with both weight loss and body fat percentage loss for subjects that are members of overweight and obese populations being treated with GLP-1RA. In addition, we evaluated whether the device-provided FLEX score is correlated with engagement with the Lumen system.

## Materials and Methods

### Study Participants

A cohort of 2296 adult Lumen (Metaflow Ltd., Tel Aviv, Israel) users who self-reported taking GLP-1RA while onboarding the Lumen smartphone application was analyzed. GLP-1RA use, as well as height, gender, and age, were ascertained via a questionnaire in Lumen’s smartphone application. It is important to note that the information about GLP-1RA use is a simple, binary “yes/no” proposition, and we lack data regarding dose, duration, and which specific GLP-1RA users take. Further information regarding age, gender, and BMI distribution of the cohort is shown in Tables 1 and 2. Weight data were collected from a combination of connected devices and manual input in the Lumen application and may be subject to measurement variability. Body fat percentage was available for only a subset of users and a combination of manually reported data and data from connected devices. Weight at 12 and 24 weeks is defined as the median weight recording through week 12 and week 24, respectively. Week 0 is defined as the first time that a new user recorded a measurement using the Lumen device. The Lumen FLEX score, a Lumen-derived estimate of metabolic flexibility, set on a scale from 7 to 21, was taken directly from the Lumen smartphone application. Details on how this parameter is quantified are found in *Supplementary Materials and Methods*.

**Table 1.**
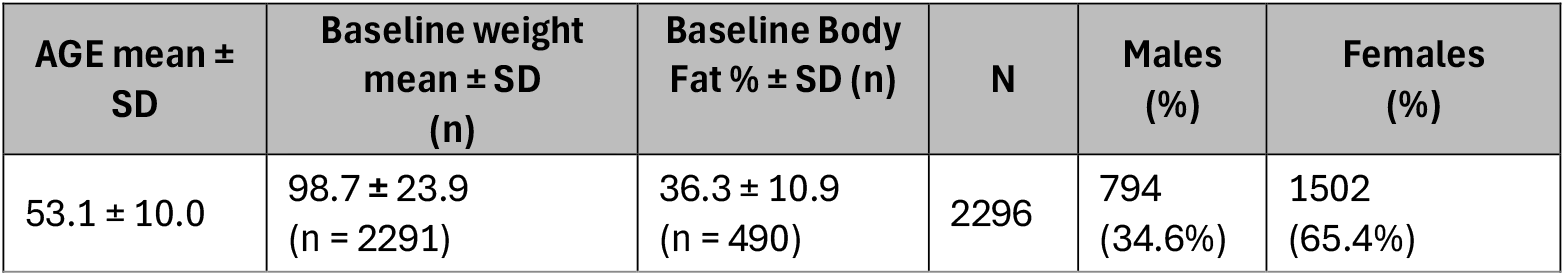
Summary of Subjects after outlier exclusion.

| AGE mean ± SD | Baseline weight mean ± SD (n) | Baseline Body Fat % ± SD (n) | N | Males (%) | Females (%) |
| --- | --- | --- | --- | --- | --- |
| 53.1 ± 10.0 | 98.7 ± 23.9 (n = 2291) | 36.3 ± 10.9 (n = 490) | 2296 | 794 (34.6%) | 1502 (65.4%) |

**Table 2.** Summary of Subjects by BMI group after outlier exclusion.

| BMI Group | AGE mean ± SD | Baseline weight mean ± SD (n) | Baseline Body Fat % ± SD (n) | N (%) | Males (%) | Females (%) |
| --- | --- | --- | --- | --- | --- | --- |
| Obese (30 and above) | 53.5 ± 10.9 | 112.2 ± 23.0 (n = 1209) | 41.8 ± 10.9 (n = 229) | 1212 (52.8%) | 445 (36.7%) | 767 (63.3%) |
| Overweight (25-30) | 53.8 ± 9.9 | 87.1 ± 13.2 (n = 748) | 32.4 ± 8.3 (n = 181) | 749 (32.6%) | 268 (36.0%) | 480 (64.0%) |
| Lean (18.5-25) | 52.6 ± 9.8 | 75.9 ± 12.0 (n = 334) | 29.2 ± 8.2 (n = 80) | 335 (14.6%) | 81 (24.2%) | 254 (75.8%) |

### Outlier Exclusion

Outliers and potentially unreliable data points were excluded via the following strategy. First, duplicate records and duplicate user IDs were excluded. All entries with heights below 100 cm or above 220 cm were excluded. Subjects with missing weight measurements or 0% change in weight throughout a user’s history were also excluded. We also excluded subjects with “implausible weight changes” as these were likely erroneous data entry or inaccurate measurement artifacts. Implausible weight changes were first flagged for manual review where the weight changed by greater than 20% in 12 weeks or greater than 35% in 24 weeks, based on clinical plausibility and prior digital weight-tracking literature. Also, subjects with underweight BMI were removed from the analysis as they are not a relevant sample when analyzing weight loss.

### Engagement Stratification

Subjects were stratified into groups of low, medium, and high engagement consistency. Criteria for each engagement consistency group are described in Table 3. Thresholds were chosen to reflect meaningful differences in weekly behavioral adherence rather than equal group sizes. A usage day is defined as a unique day on which a user used the Lumen device and smartphone application.

**Table 3.** Criteria for stratification into each engagement consistency group.

| Engagement Group | Criteria |
| --- | --- |
| High | $\geq 5$ days/week (all weeks) |
| Medium | $\geq 2$ days/week (all weeks), at least one week $< 5$ days |
| Low | at least one week $< 2$ days |

### Data Analysis

Data analysis and all statistical analysis were performed using Python. We compared outcomes across ordered engagement-consistency groups (low/medium/high) using ANCOVA adjusted for age, BMI, and sex as prespecified confounders associated with weight loss, with inference based on HC3 heteroskedasticity-robust standard errors. Model diagnostics indicated heteroscedasticity (and non-normal residuals for some outcomes) but supported a linear ordinal trend and homogeneity of slopes (no evidence of group-by-age or group-by-BMI interactions). For all comparisons of engagement consistency groups, the comparisons are ordinal (low, medium, and high engagement = 0, 1, and 2, respectively), while linear regression analyses of engagement days treat the engagement level as a continuous variable. More details regarding rationale and assumptions used in the ANCOVA analysis are found in *Supplementary Materials and Methods*.

### Ethical Considerations

This study was determined to be exempt from institutional review board (IRB) under category 2, as detailed in 45 CFR 46.104(d) and the standard operating procedure of the Biomedical Research Alliance of New York (BRANY), by the BRANY Social, Behavioral, and Educational Research IRB on May 9, 2023 (BRANY IRB File 23-119-1476). Exemption was granted because the study involved the secondary use of deidentified data that were originally collected for purposes other than this research, ensuring that participants could not be identified. All user data were fully anonymized to minimize the risk of violating participants’ privacy, and no identifiable information was accessible to the researchers. As such, informed consent was not required, and no compensation was given to the participants.

## Results

### Overall Clinical Trends

On average, the cohort of self-reported GLP-1RA users lost 3.1% of their body weight after 12 weeks and 4.4% after 24 weeks (*Figure 1A)*. They lost 1.4% of body fat percentage after 12 weeks and 2.5% after 24 weeks (*Figure 1B*).

**Figure 1.**
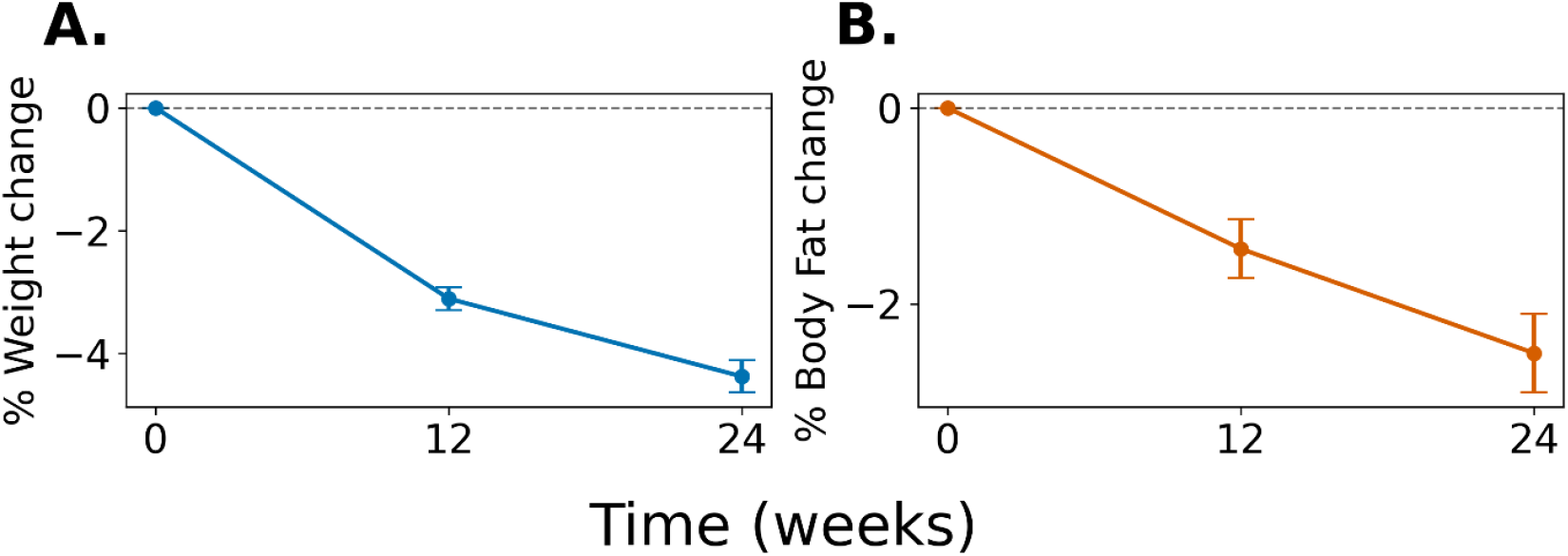
Changes in clinical parameters over time. Differences have been shown compared with time 0 in weight on a percentage basis (A) and body fat percentage (B). Plots show mean ± SEM at each timepoint.

### Influence of engagement on weight change and body fat percentage

After stratification of subjects based on their level of engagement with Lumen (stratification described in *Materials and Methods*), it became clear that weight loss was associated with increased usage of the device (*Figure 2A*), where low engagement users lost a median of 3.2% of body weight, medium engagement users lost a median of 4.6%, and high engagement users lost 5.2% of body weight. This resulted in a Spearman correlation with a p-value of 6.3×10^-11^ and a Jonckheere-Terpstra test for trends with a *P*-value of 2.4×10^-11^. This result is illustrated visually with a scatter of number of days engaged versus 24-week weight loss percentage (*Figure 2C*). As is expected in behavioral real-world data, engagement explained only a small percentage of the variability in weight change (R^2^=0.031), indicating that additional behavioral and clinical factors contribute substantially to outcomes despite a statistically detectable association (*P*(HC3)=7.8×10^− 18^).

**Figure 2.**
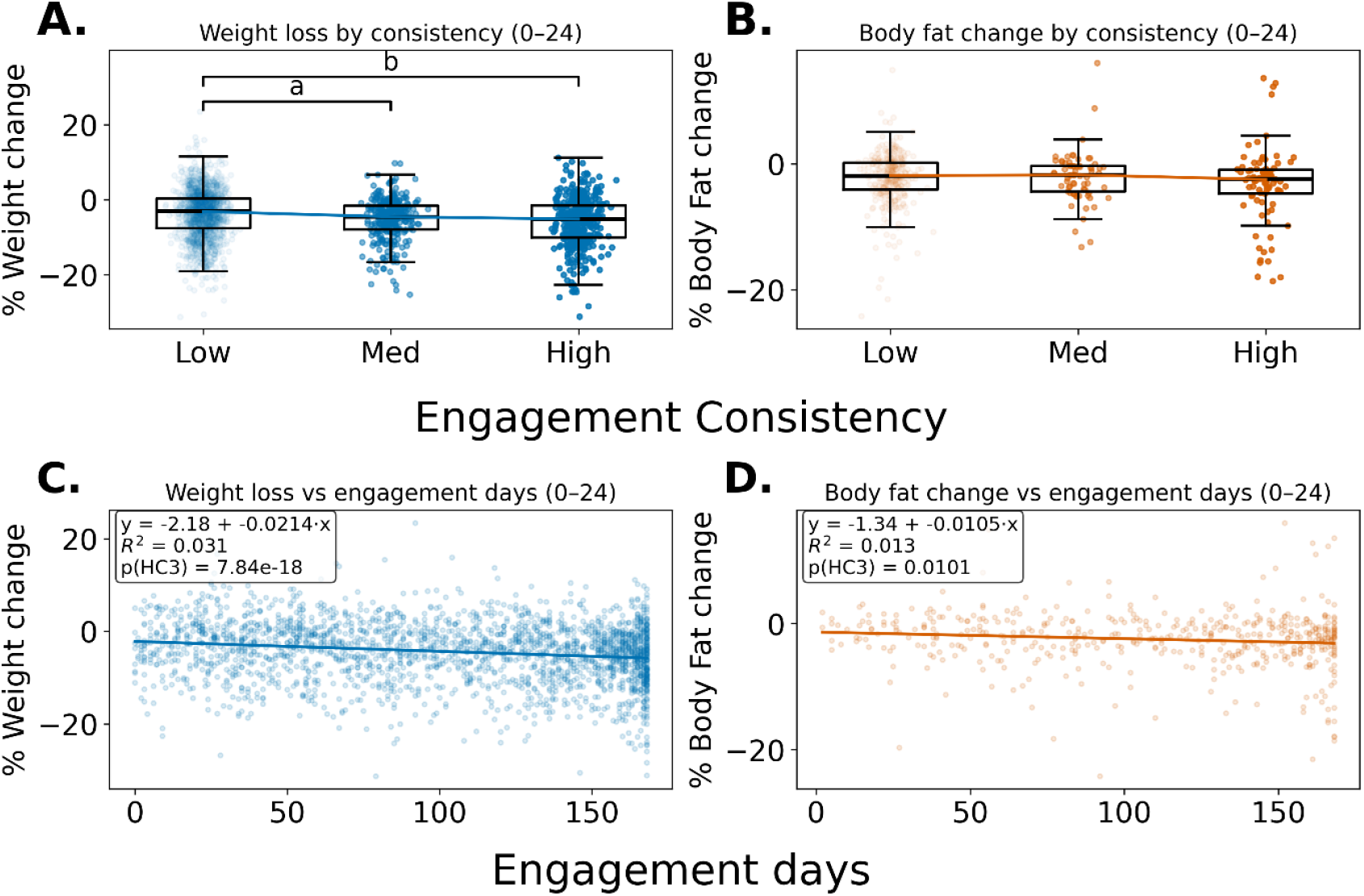
Higher engagement is correlated with more positive clinical outcomes. Engagement consistency was stratified into three groups, low (n = 1,603), medium (n = 305), and high (n = 388), and percentage change in weight (A) and percent change in body fat (B) were assessed at the 24-week timepoint. The entirety of the dataset was shown by comparing the number of days that a user engaged to each parameter (C-D). Significant P-values for the FDR-adjusted pairwise Mann-Whitney post hoc comparisons are labeled on the plots (^a^P=1.8×10^-4^; ^b^P=1.9×10^-8^).

Similar comparisons were made for body fat percentage where data were available for 490 of the 2,296 subjects in the study. Body fat percentage was also compared with stratified engagement groups (*Figure 2B*), where low engagement users lost 1.9% of body fat, medium engagement 1.8%, and high engagement 2.5%. This resulted in a Spearman correlation that did not reach statistical significance with a *P*-value of.072 and a significant Jonckheere-Terpstra test for trends with a *P*-value of.034. This result is visually illustrated with a scatter of number of days engaged versus 24-week body fat (*Figure 2D*). As is expected in behavioral real-world data, engagement explained only a small percentage of the variability in body fat percentage change (R^2^=0.013), indicating that additional behavioral and clinical factors contribute substantially to outcomes despite a statistically detectable association (*P*(HC3)=.010).

The dataset was then analyzed using ANCOVA to correct for gender, age, and initial weight/BMI. This showed a decreasing trend for weight change (*Figure 3A*) with an omnibus *P(HC3)*-value of 2.2×10^-7^. For body fat percentage change, the trend was not significant (*Figure 3B*, omnibus *P(HC3)*=.26).

**Figure 3.**
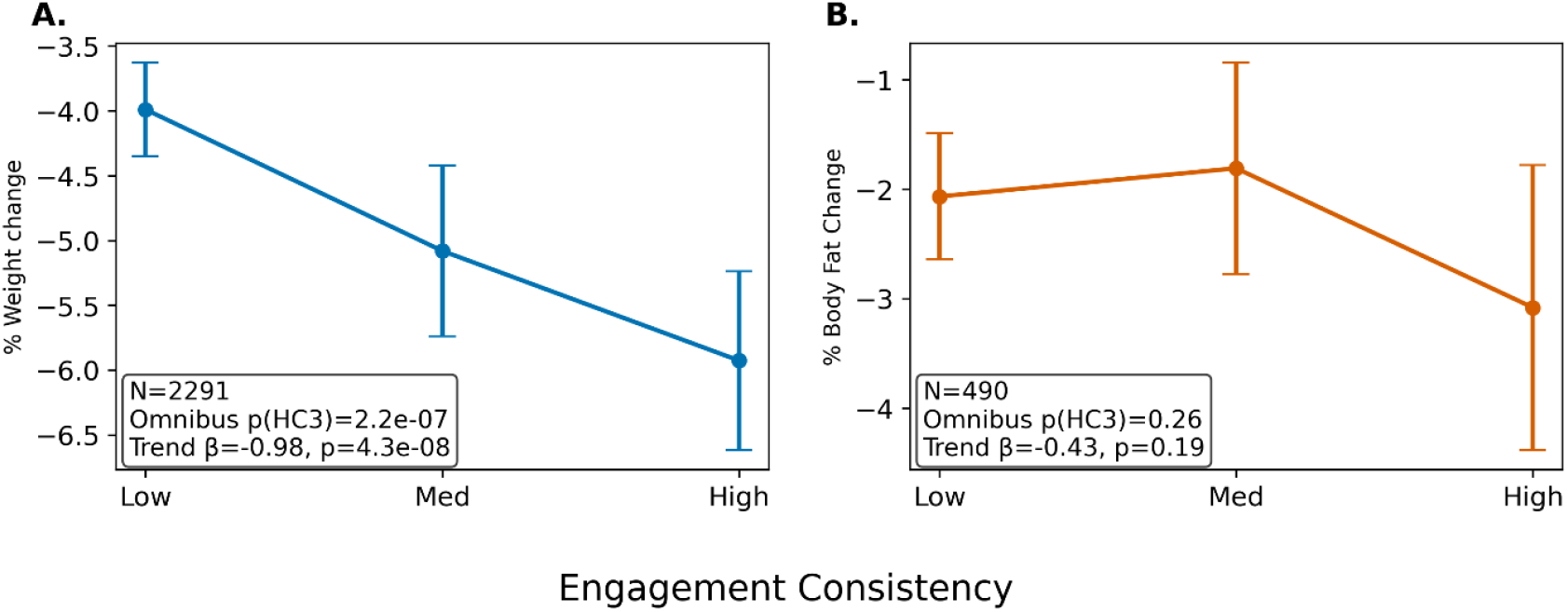
ANCOVA analysis of trends in clinical parameters. Engagement consistency was stratified into three groups, low (n = 1,603), medium (n = 305), and high (n = 388), and percentage change in weight (A) and percent change in body fat (B) were assessed at the 24-week timepoint. ANCOVA analysis was then conducted to determine whether group differences remained significant after adjustment for age, sex, and BMI

No sex-specific differences were observed in the association between engagement consistency and any outcome (all interaction *P*-values>.5, actually *P*-values are found in the *Supplementary Materials*). Stratified analyses showed consistent effects in both females and males. The association between engagement consistency and outcomes did not differ across age groups.

### FLEX Score

The FLEX score, the Lumen-derived metric for metabolic flexibility, is on a scale from 7 to 21. Across all engagement groups, after 12 weeks, the FLEX score increased by 1.08. After an additional 12 weeks, the FLEX score had decreased slightly but remained 0.90 greater than the time 0 FLEX score (*Figure 4A*). FLEX score was also compared with stratified engagement groups (*Figure 4B*), where low engagement users gained 0.08 in FLEX, medium engagement 1.7, and high engagement 2.4. This resulted in a significant Spearman correlation with a *P*-value of 6.29×10^-20^ and a significant one-sided Jonckheere-Terpstra test for trends with a *P*-value of 1.1×10^-19^. This is illustrated visually with a scatter of the number of days engaged versus 24-week FLEX score (*Figure 4C*). As is expected in behavioral real-world data, engagement explained only a small percentage of the variability in FLEX score change (R^2^=0.056), indicating that additional behavioral and clinical factors contribute substantially to outcomes despite a statistically detectable association (omnibus *P*(HC3)=2.0×10^-36^). The ANCOVA analysis for FLEX score also showed a significant increasing trend (*Figure 4D*, omnibus *P(HC3)*=4.6×10^-17^).

**Figure 4.**
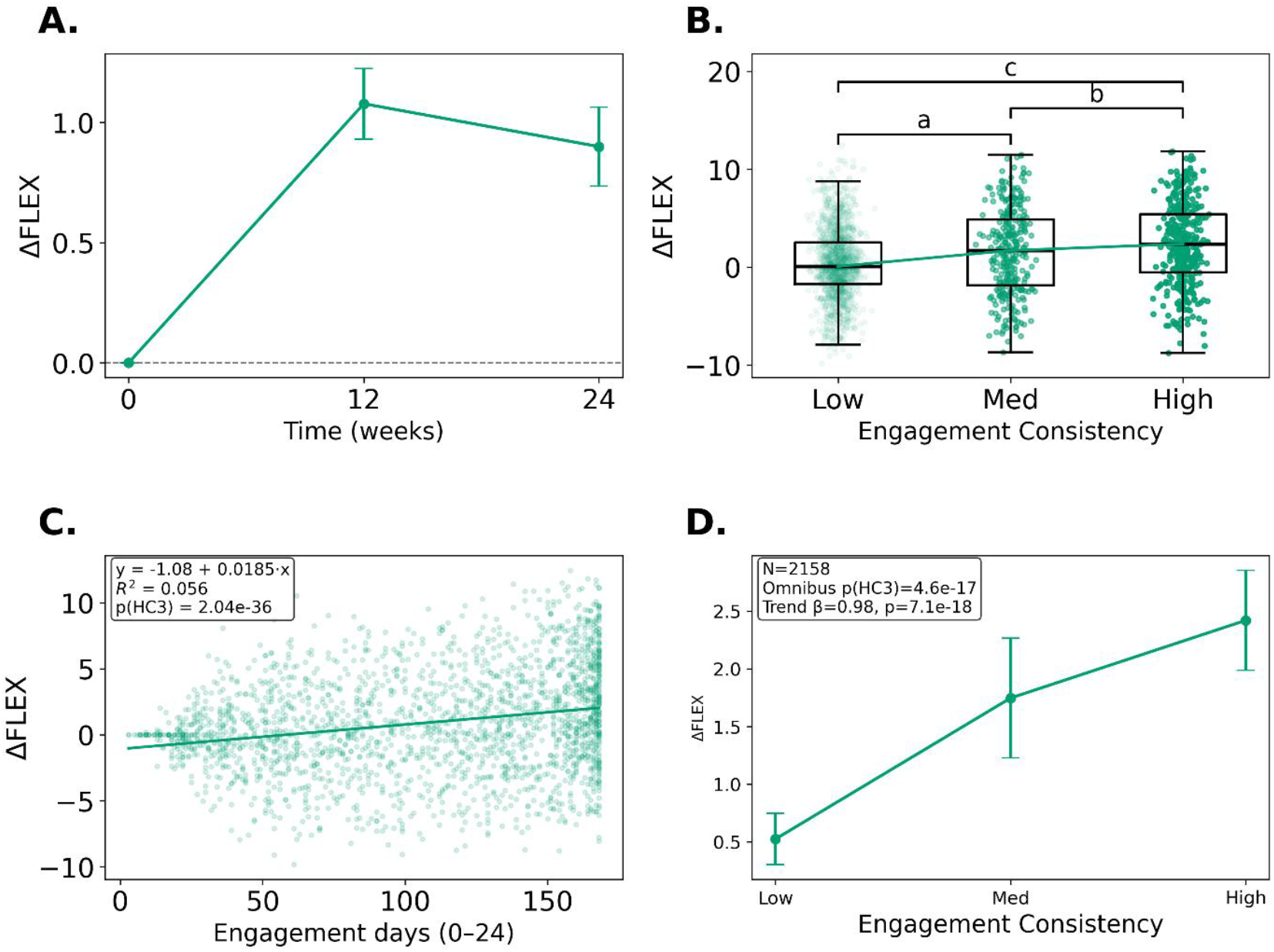
Analysis of device-derived metric intended to reflect metabolic flexibility (FLEX score). A, Differences in FLEX score over time. B, Changes in FLEX score after 24 weeks in stratified consistency groups. C, Scatter showing change in FLEX score after 24 weeks compared with number of days that a user engaged with the Lumen device. D, ANCOVA analysis of the change in FLEX score compared with engagement consistency.Significant P-values for the FDR-adjusted pairwise Mann-Whitney are labeled on the plots (^a^P=3.6×10^-6^; ^b^P=.042; ^c^P=4.1×10^-17^).

## Discussion

### Principal Results

In the present study, we have shown an association between engagement with the Lumen system and increased weight loss (3.2% weight loss for low engagement and 5.2% weight loss for high engagement). The tests for correlation between engagement and change in body fat percentage yielded mixed results. The Jonckheere-Terpstra trend test showed significance (*P*=.034), while the Spearman correlation did not reach statistical significance (*P*=.072). The fact that this correlation is weaker than that of the other parameters is confirmed both from stratifying the engagement level into low, medium, and high groups, as well as a simple correlation between number of engagement days with weight loss percentage. We also showed that Lumen’s FLEX score, a general measurement of a user’s metabolic health is correlated with both engagement and greater weight loss. ANCOVA analyses showed these associations persisted after adjustment for baseline covariates.

These findings raise the hypothesis that integrated biofeedback could augment weight loss when used alongside GLP-1RA therapy, which should be tested in controlled trials. Although engagement explains only a modest proportion of outcome variability, and causality cannot be inferred, the ∼2 percentage-point greater 24-week weight loss observed in highly engaged users (5.2% vs 3.2%) may be clinically meaningful and supports further study of whether scalable self-monitoring tools can support sustained lifestyle behaviors and improved outcomes.

### Comparison with Prior Work

We also examined whether our results match available literature regarding gender and starting weight differences in efficacy of GLP1-RA therapy. It has been reported that women show a modest but consistent advantage in weight loss compared with men [33,34]. A main explanation suggested is that women demonstrate greater engagement and adherence with diet programs [35]. Our data show no differences between male and female subjects in the correlation with engagement with the Lumen device. This pattern is consistent with hypotheses in the literature that sex differences may be mediated by behavioral factors such as adherence; however, these mechanisms were not assessed here. If this is a valid explanation, then it also follows that engagement may be more strongly associated with outcomes than gender in this cohort. The speculation that females in general show greater commitment than males is also consistent with the total cohort of GLP-1RA users in the present study being roughly two-thirds female. It should be noted that in our data, although the proportion of females increases as engagement consistency increases (65% females in the low consistency group, 66% in the medium, and 67% in the high consistency group), this is not significant. However, even the low consistency group remains a self-selected group of Lumen users and is overwhelmingly more female than the general population. It is worth noting that while it is suggested that better results among women is related to greater adherence, this is only postulated in the literature and remains unproven.

It has also been previously shown that lifestyle factors, such as diet and exercise, can significantly influence metabolic flexibility, including both acute shifts in substrate oxidation driven by recent meals and longer-term improvements associated with sustained exercise training and weight loss [13,36]. Accordingly, we hypothesized that higher engagement would be associated with higher FLEX score, which is supported by our observed associations. This is illustrated by our data that show highly significant correlation between engagement and FLEX score. FLEX should be interpreted as a device-derived metric intended to reflect metabolic flexibility and is not equivalent to metabolic-cart–derived ΔRER. It should also be noted that although the *P*-value for the interaction between FLEX and engagement is extremely strong (2.0×10^-36^), the R^2^ value is low (0.056), indicating that engagement explains only a small portion of the variation in FLEX across subjects. Most of the variability comes from other factors, such as diet composition and timing, exercise, sleep, and baseline metabolic health. It is also plausible that engagement is a marker of (or co-occurs with) differences in diet, exercise, and sleep behaviors that influence FLEX; these pathways should be examined prospectively.

### Limitations

It is important to note that this is an associative study, and we cannot make any claims regarding causality. Namely, it is evident that increased use of the Lumen device correlates with improvements in the parameters measured; however, we cannot differentiate whether the improved outcomes are the result of the metabolic information learned from the device from the possibility that more frequent use is also associated with more committed subjects, who also take diet and exercise more seriously. In addition to this issue, our GLP-1RA cohort is taken by self-response to a question in Lumen’s smartphone application and lacks detailed information such as which GLP-1RA, dose, and duration. It is entirely plausible that subjects have even discontinued GLP-1RA use during the duration of the study.

Although the improvements in weight loss and in FLEX score are highly significant, regardless of which statistical test is used or whether the engagement is stratified into groups or examined using the continuous number of usage days, the changes in body fat percentage are less significant, with the Jonckheere-Terpstra trend test yielding a significant result (*P*=.034) and the Spearman correlation not quite reaching significance (*P* =.072). There are several possible explanations for this failure to achieve significance, including the lower sample size (n=490 for body fat percentage, and n = 2291 and 2158 for weight loss and FLEX score, respectively), differences in the quality and accuracy of various measurement techniques. A future study where body fat percentage is measured in a more consistent and controlled manner would be useful, considering the concerns in the literature that GLP-1RA users lose significant lean mass in addition to fat [28,29].

### Areas for Future Study

It would be of great interest to more deeply investigate the utility of the FLEX parameter. The purpose of such a study would be to determine whether the correlation between FLEX score and weight loss is predictive or reflective. To do so, a future study could be conducted with this variable and weight loss collected at more timepoints. Then, we would examine whether the FLEX score at early timepoints is predictive of weight loss at later timepoints. If it is found to be predictive, it could be quite useful in titrating dose and personalizing therapy for GLP-1RA users. Even if it is found to be reflective rather than predictive, FLEX may have practical utility as a monitoring and feedback metric, particularly for identifying periods of waning engagement or plateau and for guiding individualized behavioral recommendations. Of course, this would need to be studied in a more controlled manner, rather than a retrospective study.

## Conclusions

In a real-world cohort of 2,296 Lumen users self-reporting GLP-1RA therapy, higher device engagement over 24 weeks was associated with greater weight loss and greater improvement in FLEX score, including after adjustment for age, baseline BMI, and sex. The association between engagement and body fat percentage reduction was smaller and did not remain significant in covariate-adjusted analyses, potentially reflecting limited body composition sample size and inconsistent measurement methods. These findings support further prospective studies to determine whether breath-based metabolic biofeedback augments GLP-1RA-associated weight loss outcomes, to clarify mechanisms, and to evaluate whether sustained engagement is associated with decreased plateauing and improved body composition trajectories. These findings should be interpreted as engagement-outcome associations rather than evidence of device efficacy.

## Data Availability

The full, noncurated dataset can be provided by request.

## Acknowledgements

The authors would like to acknowledge all app users that contributed data to this study and to the Lumen team for their support. We would also like to thank Dr. Daniel Souroujon for a thorough reading and helpful comments on the manuscript.

## Data Availability

The full, noncurated dataset can be provided by request.

## Author Contributions

MeM and MiM conceptualized the study design and supervised the study. DG and GBD led the curation of the data and its interpretation and formal analysis. RU wrote the first draft of the manuscript with significant contributions for MeM and contributed to interpretation of the data. All authors gave final approval to the manuscript.

## Abbreviations

ANCOVA: analysis of covariance
BMI: Body-mass index
BRANY: Biomedical Research Alliance of New York
CO_2_: carbon dioxide
FLEX score: Lumen-specific metric for metabolic flexibility
GLP-1RA: glucagon-like peptide-1 receptor agonist
IBT: intensive behavioral therapy

