## Supplementary Materials and Methods for "Engagement With a Breath-Based Metabolic Device Is Associated with Greater Weight Loss in Self-Reported Real-World GLP-1RA Users"

Calculation of Lumen FLEX Score

Metabolic flexibility was quantified using the Lumen FLEX score, a continuous metric computed from repeated breath-based assessments of metabolic fuel utilization. The score is designed to capture flexibility across two physiologic contexts: (1) the shift toward fat oxidation during low energy availability, assessed via app-guided morning overnight-fasted measurements, and (2) substrate switching in response to a standardized carbohydrate challenge, assessed via app-guided "boost" measurements. A boost session was defined as a structured postprandial protocol in which users consumed a high-carbohydrate meal (≥40 g carbohydrate) and then performed a breath measurement 90–120 minutes after the meal to capture the postprandial metabolic response. FLEX score was derived in two steps. First, for each calendar week, a composite score (0–100) was calculated using an algorithm based on breath-derived metabolic pattern features, including the proportion of days in fat-oxidative states and the depth of fat oxidation; when boost sessions occurred, additional weighted components reflected completion of the carbohydrate challenge and the associated postprandial response. Second, the weekly composite score was incorporated into a continuous FLEX score using exponential smoothing, with 75% weighting assigned to the prior FLEX score and 25% to the current week (after scaling the weekly composite to a fixed contribution range), thereby emphasizing sustained patterns while allowing gradual adaptation to improvement or decline. For user-facing display in the app, the resulting FLEX score was linearly transformed to a bounded range of 7–21.

ANCOVA Comparisons

For each outcome (weight change percentage, body fat percentage change, FLEX score change), we fitted ANCOVA models comparing the three ordered consistency groups (Low/Med/High; coded as CONS=1/2/3) while adjusting for age, BMI, and sex:

$$Y \sim C(CONS) + AGE + BMI + C(GENDER)$$

Inference was based on **HC3 robust standard errors** to provide reliable *P*-values and confidence intervals under potential heteroscedasticity and mild departures from OLS assumptions.

---

**Assumptions assessed and observed diagnostics (with results)**

**1) Residual normality (D'Agostino $K^2$)**

- **weight change percentage**: $P = 3.59 \times 10^{-16}$→ not met
- **body fat percentage change**: $P = 3.45 \times 10^{-19}$→ not met
- **FLEX score change**: $P = .075$→ approximately met

Given large sample sizes, normality tests are overly sensitive. We therefore treated normality as **informative** and relied on **HC3 robust inference**, rather than excluding observations or requiring strict normality.

**2) Homogeneity of variances (Levene, median-centered)**

- **weight change percentage**: $P = .0077$→ not met
- **body fat percentage change**: $P = .049$→ not met (borderline)
- **FLEX score change**: $P = 3.67 \times 10^{-13}$→ not met

Because variance heterogeneity was expected, we used **HC3 robust standard errors** to obtain valid inference under heteroscedasticity, and we report Levene's test as a diagnostic.

**3) Adequacy of a linear ordinal trend (quadratic term)**

We evaluated whether a linear trend across Low→Med→High was adequate by adding a quadratic term:

$$Y \sim CONS + CONS^2 + AGE + BMI + C(GENDER)$$

- **weight change percentage**: $P_{CONS^2} = .74$
- **body fat percentage change**: $P_{CONS^2} = .20$
- **FLEX score change**: $P_{CONS^2} = .33$

**Conclusion:** No evidence supported non-linearity; a linear ordinal trend was adequate for all three outcomes.

**4) Homogeneity of slopes (key ANCOVA assumption)**

We tested whether AGE and BMI effects differed by consistency group via an interaction model:

$$Y \sim C(CONS) * AGE + C(CONS) * BMI + C(GENDER)$$

Using **joint robust Wald tests (HC3)** on interaction terms:

- **weight change percentage**: AGE $P = .12$; BMI $P = .37$
- **body fat percentage change**: AGE $P = 0.19$ BMI $P = 0.76$
- **FLEX score change**: AGE $P = 0.43$; BMI $P = .46$

**Conclusion:** No evidence of slope heterogeneity/effect modification was detected; the homogeneity-of-slopes assumption was supported.

**5) Influential observations (leverage and Cook's distance)**

We assessed leverage and Cook's distance:

- **weight change percentage**: 120 observations with hat>2p/n; 107 with Cook>4/n; none with Cook>1
- **body fat percentage change**: 25 with hat>2p/n; 34 with Cook>4/n; none with Cook>1
- **FLEX score change**: 107 with hat>2p/n; 142 with Cook>4/n; none with Cook>1

**How handled:**

We did not automatically remove influential points. Instead, we generated a flagged-case table for transparency and potential sensitivity analyses. Importantly, no observations exceeded Cook's D>1, suggesting no extremely influential single points.

**Posthoc procedure and rationale**
We used two complementary posthoc approaches:

1. **Model-based posthoc contrasts within ANCOVA**
   Pairwise group comparisons (Med vs Low; High vs Low) were obtained as regression-based contrasts from the categorical ANCOVA model using **HC3 robust inference**. This approach is preferred over classical Tukey HSD under heteroscedasticity.
2. **Descriptive nonparametric posthoc for boxplot summaries**
   For nonparametric descriptive comparisons aligned with boxplot summaries, we conducted pairwise **Mann–Whitney U** tests (Low–Med, Med–High, Low–High) and controlled multiplicity using **Benjamini–Hochberg FDR** within each outcome.

**Summary statement**
Across outcomes, variance heterogeneity and (for weight change percentage and body fat percentage change) non-normal residuals were observed; therefore, inference relied on **HC3 robust standard errors** rather than strict parametric assumptions. Linear ordinal trend adequacy and homogeneity of slopes were supported by diagnostics. Influential observations were flagged for transparency but not automatically removed, and posthoc inference was performed via robust model-based contrasts complemented by FDR-adjusted nonparametric pairwise tests for descriptive purposes.
